# Recurrent Foodborne Outbreaks

**DOI:** 10.1101/2022.01.25.22269842

**Authors:** Sherzod B. Akhundjanov, Veronica F. Pozo, Briana Thomas

## Abstract

Firms in the food industry may experience more than one contamination incident over time. In food safety, a key objective for both the food industry and public health officials is to reduce the frequency of such incidents and extend the time between successive foodborne outbreaks. In this study, we present a systematic framework for analyzing recurrent time-to-event data, with a focus on identifying factors associated with the timing of recurrent events. Using data on meat and poultry recalls among publicly traded U.S. firms from 1994 to 2015, we find that both firm- and recall-specific characteristics are associated with overall recall risk and the risk of subsequent recalls. Recall risk increases with firm size but decreases with diversification, firm size among diversified firms, and the severity of prior recalls. Our findings highlight the importance of policies that incentivize firms to respond effectively to food safety incidents.

## 1 Introduction

Food safety is a shared responsibility across the food supply chain. Public health officials, policymakers, food firms, and consumers all play important roles in minimizing the risk of foodborne disease outbreaks.^1^ Food firms, in particular, invest in food safety technologies and protocols to prevent such incidents (Henson and Reardon, 2005). Despite these efforts, the frequency and severity of foodborne outbreaks in the United States, especially those involving meat and poultry products, remain a significant concern (Schmall, 2024). In 2015, the U.S. Department of Agriculture’s (USDA’s) Food Safety and Inspection Service (FSIS), the agency responsible for overseeing the safety of meat, poultry, and egg products, reported 150 recalls—nearly three times the number reported in 2005 (FSIS, 2017). Moreover, some firms experience multiple food safety incidents within relatively short periods. For example, Tyson Foods, one of the largest processors and marketers of chicken, beef, and pork, issued 36 separate meat and poultry recalls between 1994 and 2015 (FSIS, 2017). Tyson’s case is not unique, raising an important question: What factors influence the risk and timing of recurrent foodborne outbreaks, and how can they be properly measured?

In this study, we address this question by, first, presenting a recurrent-event survival analysis framework for repeated time-to-event settings and, second, applying it to examine how firm- and recall-specific characteristics are associated with the risk and timing of repeated food recalls. Because many food firms in our empirical setting issued more than one recall, this framework allows us to incorporate information from previous recalls when modeling the timing of subsequent events, rather than focusing exclusively on the time to a firm’s first recall, as is typical of standard survival analysis methods.

Using data on meat and poultry recalls among publicly traded U.S. firms from 1994 to 2015, we find that both firm- and recall-specific characteristics are associated with overall recall risk and the risk of subsequent recalls. Firms specializing in meat and poultry products face a higher overall recall risk as well as a higher risk of repeat recalls than more diversified firms. Both overall and repeat recall risk also increase with firm size, measured by market capitalization, although the relationship differs between diversified and less-diversified firms. Characteristics of past recalls also predict the timing of subsequent recalls. In particular, the hazard of a subsequent recall declines with the volume of product recalled in the previous incident. Firms whose previous recall was classified as Class I—the most severe category—also face a lower subsequent recall hazard than firms whose prior recall was classified as Class II. These findings highlight the importance of both firm- and recall-specific characteristics in understanding recurrent food recalls and suggest that the experience and severity of previous recall events are associated with the timing of subsequent recalls.

Our primary contribution is methodological. We demonstrate how recurrent-event survival analysis can be used to study repeated time-to-event outcomes in an economic setting. Although recurrent-event methods are well established in epidemiology and biostatistics, where they have been used to study outcomes such as bladder tumor recurrence (Amorim and Cai, 2015), cancer recurrence (Rondeau et al., 2007; Sultana et al., 2026), recurrent asthmatic events (Duchateau et al., 2003), repeated acute lower respiratory infections (Kelly and Lim, 2000; Amorim and Cai, 2015), and hospital readmissions among the elderly (Kennedy et al., 2001), their adoption and use in economics has been more limited. As discussed below, conventional approaches to repeated events often focus either on the time to a single event or on the total number (count) of events experienced by an observational unit. Neither approach fully exploits the timing and ordering of repeated events. Recurrent-event survival analysis, by contrast, is specifically designed to model the timing of multiple events experienced by the same unit. The paper evaluates alternative approaches for modeling recurrent time-to-event data and examines the implications of alternative model specifications for the analysis and interpretation of recurrent events. We apply this framework to food recalls (agricultural economics), but the approach is applicable to other economic settings characterized by recurrent events, including repeated pharmaceutical product recalls (health economics), recurrent consumer, durable, and non-durable product recalls (industrial organization), and repeated environmental violations (environmental economics). The food-safety application therefore serves as an empirical setting in which to illustrate the broader utility of recurrent-event methods for economic research.

Our empirical findings also have managerial and policy implications. In food safety, the time between successive recall events provides an important measure of a firm’s ability to prevent recurrent safety incidents. Identifying the firm and recall characteristics associated with this interval can provide managers with information relevant to the design of food safety protocols and quality management systems (e.g., acceptance sampling and statistical process control). Reducing the frequency of recalls may also help firms avoid the economic costs associated with product withdrawals and disruptions while improving consumer food safety. From a policy perspective, the lower subsequent recall hazard associated with more severe prior recalls is consistent with the possibility that firms respond to particularly consequential food safety incidents (i.e., Class I recalls) by strengthening their safety practices. At the same time, the heterogeneity in recall risk across firms and across prior recall characteristics suggests that inspection and enforcement programs may benefit from accounting for differences in firms’ histories of food safety incidents.

The remainder of the article is organized as follows. The next section provides background and reviews the related literature. Section 3.1 presents the data, and Section 3.2 describes the empirical methodology used to analyze recurrent food recalls. Section 4 reports the results. Section 5 concludes.

## 2 Background and Literature

Food recalls occur when a contaminated food product has been distributed to the market and, depending on the severity of the contamination, may pose serious health hazards.^2^ In the United States, firms are responsible for removing recalled products from the marketplace, subject to oversight by federal or state authorities (FDA, 2017).^3^ For meat and poultry products, the FSIS issues a public recall announcement specifying the firm and product involved, the reason for the recall, the severity of the potential health threat (i.e., recall class), and the quantity of product recalled. The FSIS classifies food recalls into three broad categories. Class I recalls are the most serious and involve a reasonable probability that consumption of the product will cause adverse health consequences or death. Class II recalls involve a remote probability of adverse health consequences, whereas Class III recalls involve products that are not expected to cause adverse health consequences.

Recalls can impose substantial costs on firms, including the direct costs of removing products from the market as well as litigation expenses, reputational damage, and reduced stakeholder confidence (Jarrell and Peltzman, 1985; Pruitt and Peterson, 1986; Welling, 1991; Sockett, 1993; Rhee and Haunschild, 2006; Chen et al., 2009; Hua, 2011). Accordingly, much of the existing literature has focused on quantifying the direct and/or indirect economic costs of food recalls (Henson and Mazzocchi, 2002; Marsh et al., 2004; Piggott and Marsh, 2004; McCluskey et al., 2005; Mazzocchi, 2006; Schroeder et al., 2007; Spalding et al., 2023). Quantifying these costs is important for firms seeking to evaluate the economic consequences of food-safety failures and design food-safety strategies. From a policy perspective, however, it is also important to understand why some firms experience repeated food-safety incidents more frequently than others. Identifying the factors associated with the risk and timing of recurrent recalls can provide additional insight into the determinants of persistent food-safety problems.

The literature examining the determinants of repeated food recalls remains limited (Teratanavat et al., 2005; Hall and Johnson-Hall, 2017). Existing studies typically employ either standard survival models or panel generalized linear models (GLMs), such as negative binomial specifications. Conventional survival models are designed primarily to characterize the time to a single event (e.g., a firm’s first recall) and therefore do not directly account for the recurrent occurrence of events within firms (Kleinbaum and Klein, 2005; Amorim and Cai, 2015). Panel count models, in contrast, are well suited to modeling the number of recalls experienced by a firm over a given period but do not model the timing of individual recall events. Thus, neither approach fully captures the recurrent time-to-event nature of food recalls.

To our knowledge, Akhundjanov et al. (2024) is the only other study that applies a recurrent-event survival analysis framework to repeated compliance failures in an economic setting. That study focuses mainly on identifying evidence of learning following repeated compliance issues, whereas we examine factors associated with the risk and timing of recurrent food recalls. This difference in research objectives also motivates a methodological distinction. Akhundjanov et al. (2024) employ the Prentice-Williams-Peterson Gap-Time (PWP-GT) stratified Cox proportional hazards model (Prentice et al., 1981), which accounts for the ordering of recurrent events and models the time elapsed between successive events. We instead consider three complementary recurrent-event regression approaches—the Andersen-Gill (AG) counting-process model (Andersen and Gill, 1982), the Prentice-Williams-Peterson Gap-Time (PWP-GT) model, and frailty models—together with Kaplan-Meier survival curves (Kaplan and Meier, 1958) to characterize the determinants and timing of recurrent recalls. Taken together, the two studies illustrate the utility of recurrent-event methods for analyzing repeated time-to-event outcomes in economics and provide complementary approaches for studying repeated compliance and food-safety failures.

## 3 Methods

### 3.1 Data

This study uses data from the USDA FSIS Recall Case Archive, covering meat and poultry recalls issued by publicly traded firms in the United States between 1994 and 2015.^4^ The FSIS provides information on the firm issuing the recall, the recall date, the quantity of product recalled, and the recall classification. Table 1 presents the number of recalls and the total quantity recalled for the 31 publicly traded firms in our sample. The sample includes many major meat and poultry processors and marketers and encompasses considerable heterogeneity across firms. Some firms highly specialized, such as Sanderson Farms, which primarily produces raw chicken products, whereas others, such as Kraft, operate across multiple food-product categories. The firms also vary substantially in market capitalization and age.

**Table 1:**
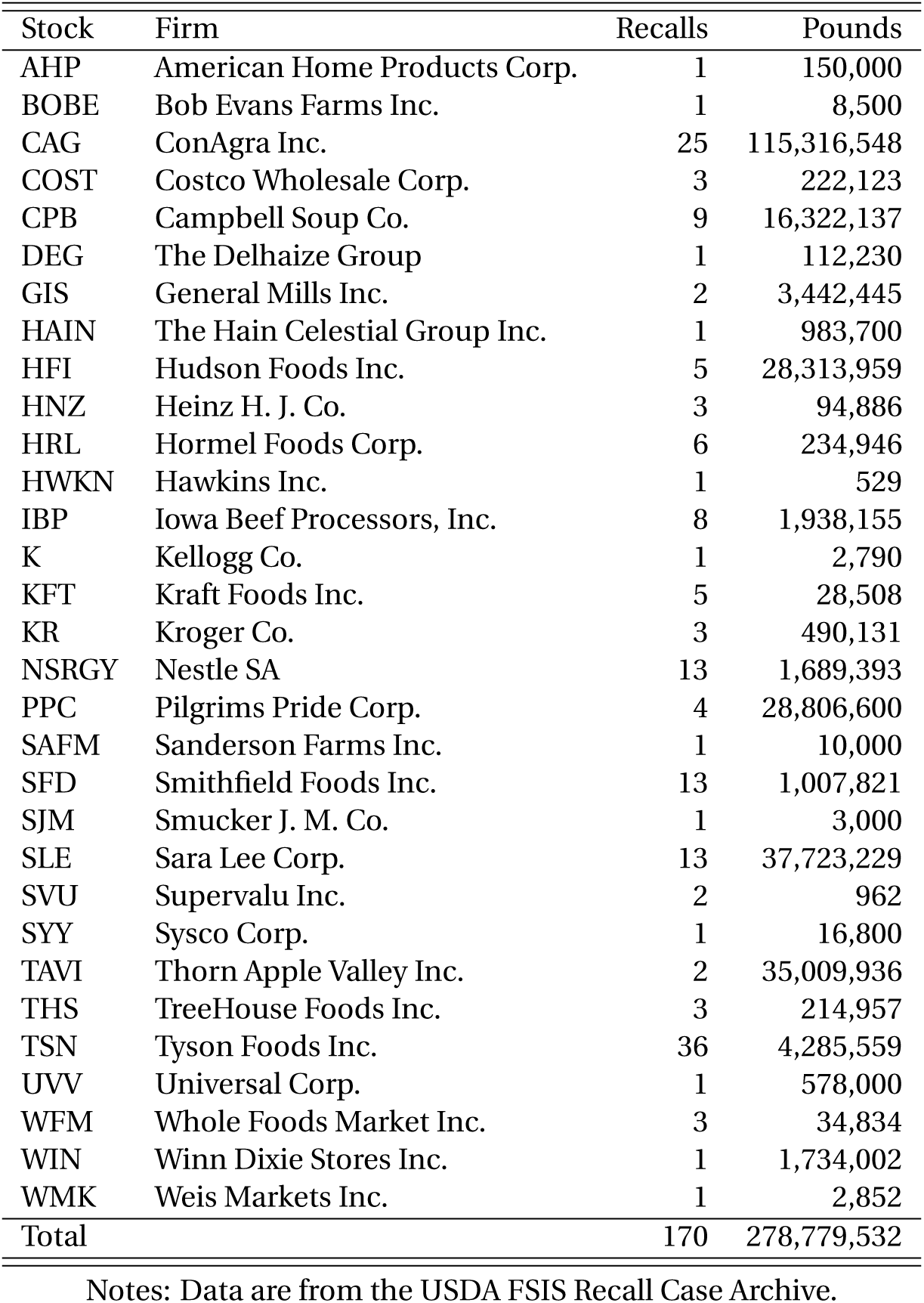
Number of recalls and quantity of meat and poultry recalled by publicly traded firms in the United States, 1994-2015.

In the recurrent-event framework, each firm constitutes a unit and each recall constitutes an event. Since the event can recur, these events are termed recurrent events. We define a firm’s entry into the risk set as January 1, 1994, the date on which the FSIS began collecting recall data, or the date of the firm’s initial public offering (IPO), whichever occurs later. This definition ensures that the time to the first event is measured only over the period during which the firm is observed in the sample. The observation period ends in December 2015. Firms that were acquired or filed for bankruptcy before the end of the study period are right-censored at the date of acquisition or bankruptcy filing.^5^ The resulting sample contains 201 firm-duration observations corresponding to 170 recall events.

Our key variable, *Duration till Recall*, measures the elapsed time (in months) from entry into the risk set or the preceding recall to the next recall or censoring. To account for the ordering of recurrent recall events, we stratify duration observations by event interval (stratum). Stratum 1 represents the time to the first recall event, stratum 2 the time from the first to the second recall, and so forth. Because the maximum number of recalls experienced by a firm in our sample is 36, the final stratum (stratum 37) represents the time from the 36th recall to the end of the observation period or censoring.

We consider three firm-level characteristics: firm size, whether meat and poultry constitute the firm’s primary output, and firm age. We construct these measures using information from firms’ annual reports and Form 10-K filings. *Firm Size* is measured by market capitalization (in millions of U.S. dollars), calculated as the number of shares outstanding multiplied by the stock price observed 10 days before the recall announcement (Fama and French, 1992). Market capitalization is time-varying and therefore captures changes in firm size over the sample period. We deflate market capitalization using the Consumer Price Index (CPI) from the U.S. Bureau of Labor Statistics.^6^ *Meat Main* is an indicator equal to one when meat or poultry products constitute the firm’s primary output and zero otherwise. This variable distinguishes primarily meat and poultry producers from diversified (multi-product) food manufacturers and retailers. *Firm Age* is also time-varying and measures the number of years since the firm’s establishment.

While controlling for other firm-specific characteristics, such as product-specific production levels and investments in food-safety technologies, would be desirable, such information is generally limited or unavailable.^7^ Our firm-level controls therefore provide indirect measures of these characteristics. In particular, firm size may capture differences in production capacity and food-safety practices. Evidence from the Economic Research Service (ERS) indicates that larger food firms tend to have greater sanitation, process-control, and laboratory capabilities (Ollinger and Mueller, 2003) and invest more in sanitation equipment and testing technologies (Ollinger et al., 2004). Firm age may similarly capture accumulated organizational experience. The set of firm-level controls is comparable to those used in previous empirical studies of food recalls (Henson and Mazzocchi, 2002; Teratanavat and Hooker, 2004; Teratanavat et al., 2005; Spalding et al., 2023; Akhundjanov et al., 2024). Nevertheless, our methodology accounts for remaining unobserved firm-level heterogeneity through a shared frailty specification (see Section 3.2.3).

We also consider two recall-specific characteristics: recall class and recall size. We obtain both measures from the USDA FSIS Recall Case Archive. We construct three mutually exclusive indicators for recall severity: *Class I*, *Class II*, and *Class III*, each equal to one when the corresponding recall classification applies and zero otherwise. *Recall Size* measures the total quantity of product recalled (in thousands of pounds) in a given event.

Table 2 reports descriptive statistics, and Figure 1 presents the corresponding data visualizations. All 31 firms experienced at least one recall during the study period. Recall frequency varies substantially across firms: approximately half experienced three recalls, five experienced 13 recalls, two experienced 25 recalls, and one experienced 36 recalls (Table 1 and Figure 1(a)). On average, firms in the sample experienced about eight recall events (Table 2). Class I recalls constitute the largest share of recalls, followed by Class II and Class III recalls (Table 2). Although diversified firms account for a larger share of recalls within each event stratum (Figure 1(b)), firms whose primary output is meat or poultry, which constitute 38% of the sample (Table 2), experience more recalls per firm. The distribution of duration times also varies across event strata (Figure 1(c))). The time to the first recall (stratum 1) is generally longer than the intervals between subsequent recalls, with stratum 2 exhibiting the next-longest duration. Duration declines from stratum 2 to stratum 3, increases in stratum 4, and then remains relatively stable across subsequent strata. The bivariate relationships between duration and firm- and recall-specific characteristics are less pronounced (Figure 1(d-f)), although duration appears negatively associated with firm size. We examine these relationships formally using recurrent-event survival models.

**Figure 1:**
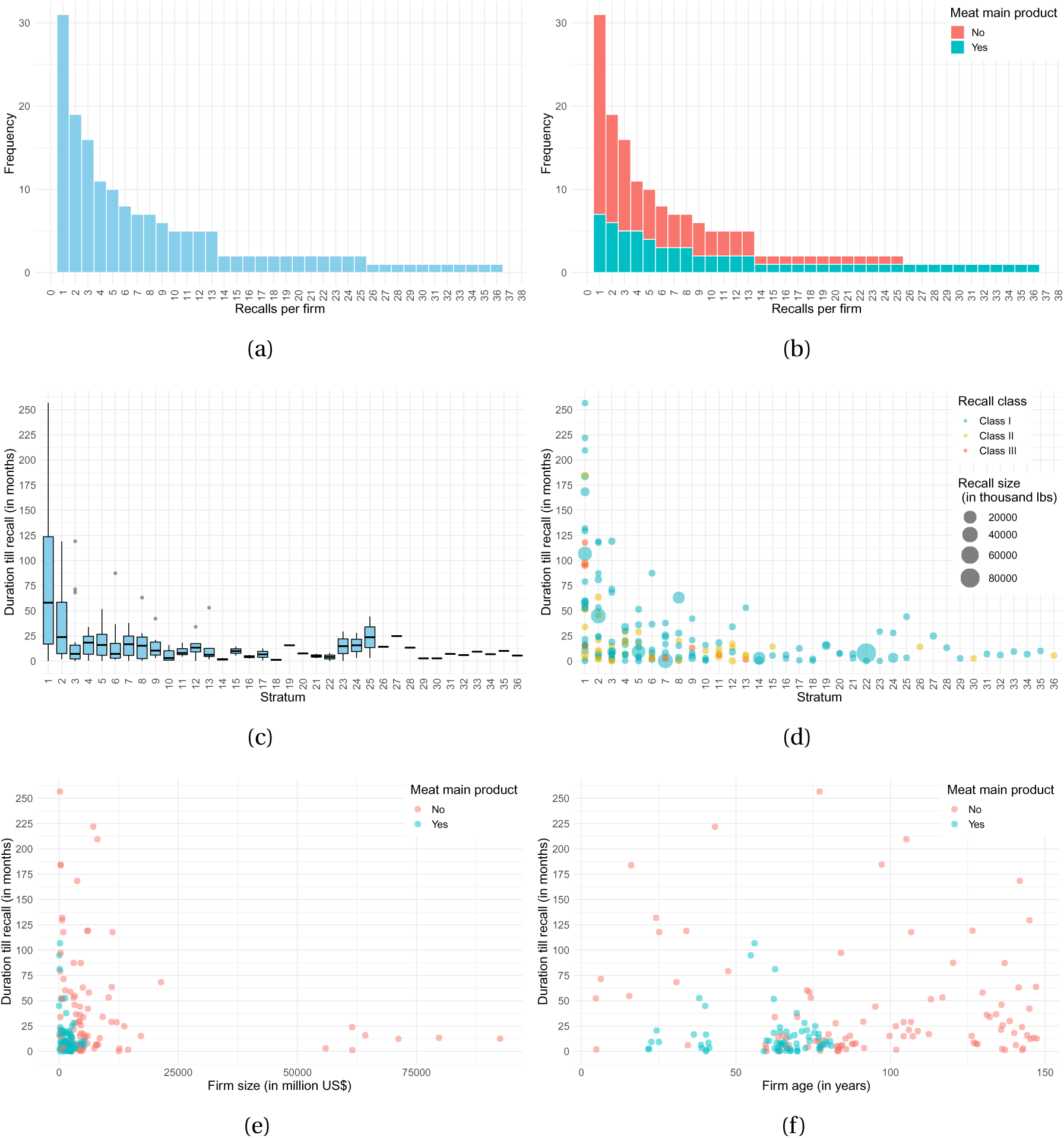
Characteristics of study recalls and firms issuing them. Notes: The sample consists of meat and poultry recalls issued by publicly traded firms in the United States between 1994 and 2015. Data are from the USDA FSIS Recall Case Archive.

**Table 2:**
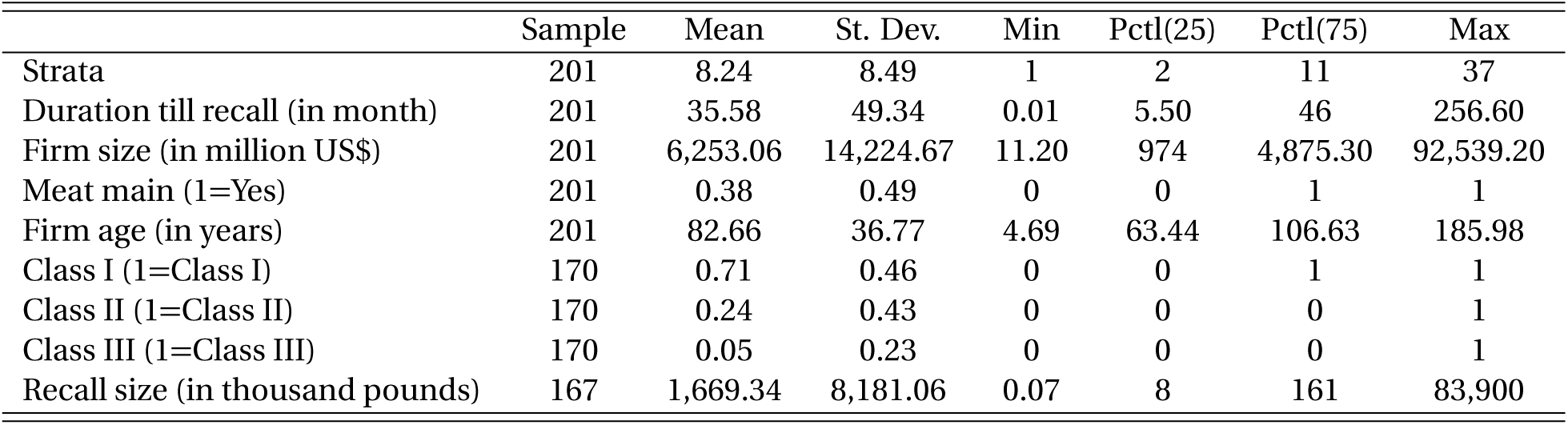
Descriptive statistics.

### 3.2 Statistical Analysis

Because the outcome of interest may occur repeatedly for the same unit, we model the timing of recurrent events using three extensions of the Cox proportional-hazards framework: the Andersen-Gill (AG) counting-process model (Andersen and Gill, 1982), the Prentice-Williams-Peterson Gap-Time (PWP-GT) model (Prentice et al., 1981), and frailty models. All three models accommodate right censoring and time-varying covariates, but they differ in how they characterize the recurrent-event process, including the treatment of event history, risk-set construction, and unobserved unit-specific heterogeneity (Kelly and Lim, 2000; Kleinbaum and Klein, 2005; Amorim and Cai, 2015). These distinctions are important because the three specifications characterize somewhat different aspects of the process generating repeated events.

#### 3.2.1 Andersen-Gill Model

The counting process model used in this study was developed by Andersen and Gill (1982) and is commonly referred to as the Andersen-Gill (AG) model. The AG specification can be viewed as the recurrent-event analogue of the standard Cox proportional hazards model (Cox, 1972), with the event history incorporated through the counting-process representation. A unit remains in the risk set after experiencing an event, provided that it remains under observation and at risk of experiencing another event.

The AG model specifies the hazard or intensity for unit *i* during the *k*th event interval (stratum) as

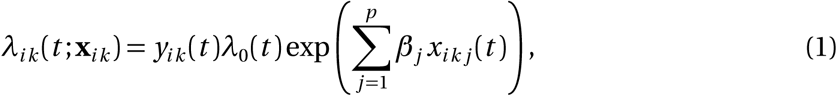

where *t* denotes the observed duration until an event or censoring, **x***_ik_* (*t*) = (*x_ik_*_1_, …, *x_ikp_*)*^′^* is a vector of possibly time-varying covariates, ***β*** = (*β*_1_, …, *β_p_*)*^′^* is a vector of unknown coefficients, *y_ik_* (*t*) indicate whether unit *i* is at risk of experiencing an event at *t*, and *λ*_0_(*t*) is an unspecified baseline hazard corresponding to **x***_ik_* (*t*) = **0**.

The AG model has several features that merit explicit discussion.

##### Common baseline hazard

The AG model treats recurrent events as realizations of a single counting process observed over the study’s total-time scale and specifies a common baseline hazard across recurrent events. Hence, the same baseline hazard *λ*_0_(*t*) applies to the first, second, and subsequent events. Event order therefore does not enter the baseline hazard directly. Consequently, the AG model coefficients characterize an *overall association*between the observed covariates and the hazard of recurrent events over time rather than an association specific to a particular event order.

If event order is expected to affect the baseline hazard beyond what is captured by the observed covariates, conditional recurrent-event models, such as the Prentice-Williams-Peterson Gap-Time (PWP-GT) model, provide an alternative. As discussed below, the PWP-GT model conditions the risk set on the occurrence of previous events and allows the baseline hazard to vary across event orders, or strata (Kleinbaum and Klein, 2005).

##### Proportional hazards

Conditional on the observed event history, exp(*β_j_*) is the multiplicative effect of a one-unit increase in *x_ik j_* (*t*) on the instantaneous recurrent-event hazard. Thus, the AG model assumes that the relative effect of *x_ik j_* (*t*) on the event hazard is proportional over time. The proportional hazards (PH) assumption can be assessed using tests based on Schoenfeld residuals (Grambsch and Therneau, 1994). Evidence against the PH assumption indicates that the covariate effect varies over time and may motivate a more flexible specification, such as interactions between covariates and event strata.

##### Conditional independence

The AG model assumes that, conditional on the observed covariates and event history, there is no additional systematic dependence in the recurrent-event process attributable to unobserved unit-specific heterogeneity. The AG model is therefore most appropriate when dependence among recurrent events can be adequately characterized by observed covariates and event history rather than by unobserved unit-specific heterogeneity. In our empirical setting, the conditional independence assumption may have some regulatory support. When a firm issues a recall, the FSIS closely monitors the recall process. If it is subsequently determined that more product is affected than initially reported, a recall extension is issued. Hence, when an issue associated with an existing recall persists, it is generally addressed through an extension of the original recall rather than through a new recall (FSIS, 2013). This regulatory practice reduces the likelihood that multiple observations associated with the same underlying incident are recorded as separate recall events and supports treating observed recalls as distinct events in our recurrent-event framework.

If within-unit dependence remains after conditioning on the observed covariates and event history, potentially because units differ in unobserved characteristics that persist over time, two approaches can be considered. First, a unit-level robust (sandwich) variance estimator can be used for inference (see Amorim and Cai, 2015), as implemented in all of our AG analyses. This estimator accommodates within-unit dependence in the estimating equations, without specifying the source of that dependence, and provides robust variance estimates under general forms of within-unit correlation. It does not, however, correct misspecification of the conditional hazard or the common-baseline restriction. Second, unobserved unit-specific heterogeneity can be modeled explicitly using frailty models, which we discuss below.

Separate from the considerations discussed above, because multiple units may experience events at the same observed time, the Breslow approximation is used to account for tied failure times (Breslow, 1974).

#### 3.2.2 Prentice-Williams-Peterson Gap-Time Model

The conditional recurrent-event model used in this study was developed by Prentice et al. (1981) and is commonly referred to as the Prentice-Williams-Peterson (PWP) model. The PWP model can be formulated as a stratified Cox proportional hazards model in which the strata correspond to the order of recurrent events. Two versions of the PWP model have been proposed. The first, known as conditional 1 or the PWP Total-Time (PWP-TT) model, uses time from study entry as the underlying time scale. The second, known as conditional 2 or the PWP Gap-Time (PWP-GT) model, uses the time elapsed since the previous event as the underlying time scale. We adopt the PWP-GT model because our primary interest is in the time to the next recall following a previous recall, rather than in time measured from study entry to the first, second, and subsequent recalls (Kleinbaum and Klein, 2005). Our implementation follows that of the companion paper by Akhundjanov et al. (2024).

Unlike the AG model, the PWP-GT model explicitly accounts for event order by stratifying the risk sets according to the number of previously experienced events. The risk set for the *k* th event consists only of units that have already experienced their (*k* − 1)st event. Hence, a unit cannot contribute to the risk set for the *k* th event unless it has first experienced the preceding event. This stratification allows the baseline hazard to vary across event orders and, when covariate-by-stratum interactions are included, permits the effects of covariates to differ across recurrent recalls (Kelly and Lim, 2000; Kleinbaum and Klein, 2005; Amorim and Cai, 2015). In a comparison of alternative recurrent-event models, including AG, PWP-TT, and PWP-GT, Kelly and Lim (2000) highlight the utility of the PWP-GT model for analyzing recurrent-event data.

The PWP-GT model specifies the hazard or intensity for unit *i* during *k*th event interval (stratum) as

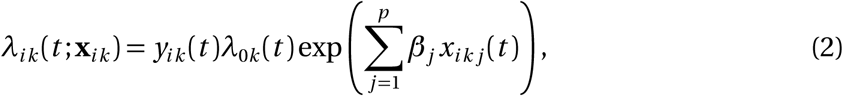

where *λ*_0*k*_ (*t*) is the stratum-specific baseline hazard. The underlying time scale *t* is the elapsed time since the previous recall. The event-specific baseline hazards allow the baseline risk of a subsequent recall to vary across event orders. This feature is important when the occurrence of previous recalls is itself informative about the timing of subsequent recalls and distinguishes the PWP-GT model from the AG model, which imposes a common baseline hazard across event orders.

The PWP-GT model coefficients characterize the association between the observed covariates and the hazard of the *next event*, conditional on the occurrence of the preceding event. This gap-time formulation is well suited to settings in which the interest lies in the duration between successive events rather than in the cumulative occurrence of events over study time.

##### Proportional hazards

Similar to the AG model, the PWP-GT specification imposes the PH assumption, under which the effects of observed covariates are multiplicative and remain constant over the relevant gap-time scale. The PH assumption can be evaluated using tests based on Schoenfeld residuals (Grambsch and Therneau, 1994).

##### Conditional independence

The PWP-GT specification also assumes that, conditional on the observed covariates and event history, there is no additional systematic dependence in the recurrent-event process attributable to unobserved unit-specific heterogeneity. To account for any remaining within-unit dependence in statistical inference, a unit-level robust (sandwich) variance estimator can be used (see Amorim and Cai, 2015). Alternatively, unobserved unit-specific heterogeneity can be modeled explicitly using frailty models, as discussed next.

Separately, because events may occur at the same observed time for multiple units, the Breslow approximation is implemented to account for tied failure times (Breslow, 1974).

#### 3.2.3 Frailty Models

Although the AG and PWP-GT models account for the recurrent nature of events, neither model explicitly introduces a unit-specific source of unobserved heterogeneity. This consideration becomes potentially important if units differ in unobserved characteristics that persist over time and affect their propensity to experience events. In our empirical setting, for example, firms may differ in food-safety practices, production processes, quality-control systems, managerial practices, or the effectiveness of internal monitoring. If such characteristics are not observed, recalls generated by the same firm may remain correlated even after conditioning on the observed covariates and event history.

To accommodate this possibility, we consider a shared frailty model (Amorim and Cai, 2015; Balan and Putter, 2020). The frailty model introduces an unobserved unit-specific random effect that captures persistent heterogeneity not explained by the observed covariates and induces dependence among recurrent event times. Let *u_i_* denote the unit-specific random effect (frailty), assumed to follow a gamma distribution with mean one, E(*u_i_*) = 1, and variance *θ*, Var(*u_i_*) = *θ*. The conditional hazard or intensity for unit *i* during the *k*th event interval (stratum) is specified as

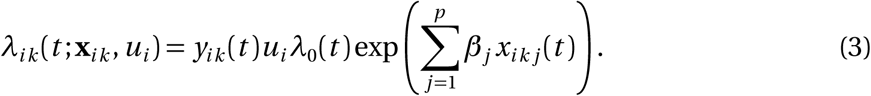

The frailty *u_i_* is assumed to be constant for a given unit over the observation period, thereby capturing persistent unobserved heterogeneity that affects the occurrence of recurrent events. Units with relatively high frailty *u_i_* tend to experience events more frequently, whereas units with relatively low frailty tend to experience them less frequently. Frailty thus provides an explicit representation of within-unit dependence arising from persistent unobserved heterogeneity.

The frailty model coefficients in (3) characterize an *overall association* between the observed covariates and the hazard of recurrent events over time, conditional on firm-specific frailty (random effects). This interpretation is similar to that of the AG model, with the important distinction that the frailty model explicitly accounts for persistent unobserved heterogeneity across units. Conditional on the unit-specific frailty and observed covariates, recurrent events are assumed to be independent.

The frailty model also provides a useful diagnostic for the importance of unobserved heterogeneity. The frailty variance, Var(*u_i_*) = *θ*, measures the degree of persistent unobserved heterogeneity across units after conditioning on the observed covariates. If *θ* = 0, all units have the same frailty, and the model reduces to the corresponding Cox proportional hazards model (e.g., AG or PWP-GT model). As *θ* increases, unobserved heterogeneity becomes more pronounced and induces stronger dependence among recurrent events within units under the shared-frailty specification.

##### Proportional hazards

The PH assumption remains relevant: conditional on frailty, the effect of the observed covariates is assumed to be multiplicative and constant over the relevant time scale. Introducing frailty therefore does not by itself resolve violations of PH. Rather, its purpose is to account for persistent unobserved heterogeneity that generates dependence among recurrent events. The PH assumption can be tested using Schoenfeld residuals (Grambsch and Therneau, 1994).

As a final technical note, the Breslow approximation is employed to account for tied failure times (Breslow, 1974).

## 4 Results

For our application, the three models provide complementary approaches to modeling recurrent recalls. The AG and frailty models focus on the *overall* recall risk over study time, whereas the PWP-GT model focuses on the risk of the *next recall*. Comparing these specifications allows us to evaluate whether the estimated associations between the observed covariates and recall timing are sensitive to alternative assumptions about event history and unobserved firm-level heterogeneity.

Recurrent-event survival analysis is notoriously sensitive to the number of events observed in each stratum because the number of events in later strata may become small (e.g., Table 1), affecting the stability and precision of the estimates (Kelly and Lim, 2000; Kleinbaum and Klein, 2005; Amorim and Cai, 2015). We therefore conduct the analysis using two samples: (i) a restricted sample containing the first eight event strata, corresponding to the mean number of recall events per firm (Table 2), and (ii) the full sample containing all observed event strata. This comparison allows us to check whether the results are sensitive to the inclusion of sparsely populated higher-order event strata.

### 4.1 Factors Associated with the Timing of Recurrent Food Recalls

Table 3 reports the estimation results from the recurrent-event survival analysis based on the first eight recall events, corresponding to the mean number of recalls per firm. Across alternative specifications of the observed covariates, the estimates from the AG, PWP-GT, and frailty models are largely consistent, both quantitatively and qualitatively. The results suggest that *Firm Age* is not statistically significantly associated with recall risk, either in terms of overall recall risk or the risk of subsequent recalls.

**Table 3:** Estimates from recurrent-event survival models: first eight strata.

|  | AG<br>(1) | PWP-GT<br>(2) | Frailty<br>(3) | AG<br>(4) | PWP-GT<br>(5) | Frailty<br>(6) | AG<br>(7) | PWP-GT<br>(8) | Frailty<br>(9) |
| --- | --- | --- | --- | --- | --- | --- | --- | --- | --- |
| Firm size | 4.83×10<sup>-5***</sup><br>(1.12×10<sup>-5</sup>)<br>[1.00] | 2.16×10<sup>-5**</sup><br>(9.78×10<sup>-6</sup>)<br>[1.00] | 2.96×10<sup>-5**</sup><br>(1.48×10<sup>-5</sup>)<br>[1.00] | 4.66×10<sup>-5***</sup><br>(1.00×10<sup>-5</sup>)<br>[1.00] | 2.39×10<sup>-5**</sup><br>(9.98×10<sup>-6</sup>)<br>[1.00] | 3.20×10<sup>-5**</sup><br>(1.48×10<sup>-5</sup>)<br>[1.00] | 4.04×10<sup>-5***</sup><br>(1.05×10<sup>-5</sup>)<br>[1.00] | 1.64×10<sup>-5*</sup><br>(9.53×10<sup>-6</sup>)<br>[1.00] | 3.77×10<sup>-5**</sup><br>(1.48×10<sup>-5</sup>)<br>[1.00] |
| Meat main | 1.08***<br>(0.39)<br>[2.95] | 0.78**<br>(0.31)<br>[2.18] | 1.07**<br>(0.51)<br>[2.93] | 0.45<br>(0.40)<br>[1.58] | 0.47<br>(0.40)<br>[1.60] | 0.56<br>(0.58)<br>[1.74] | 1.07***<br>(0.37)<br>[2.92] | 0.92***<br>(0.34)<br>[2.50] | 1.04**<br>(0.41)<br>[2.83] |
| Firm age | -7.97×10<sup>-4</sup><br>(3.10×10<sup>-3</sup>)<br>[1.00] | -2.88×10<sup>-3</sup><br>(2.66×10<sup>-3</sup>)<br>[0.99] | -6.59×10<sup>-3</sup><br>(5.22×10<sup>-3</sup>)<br>[0.99] | -9.31×10<sup>-4</sup><br>(3.07×10<sup>-3</sup>)<br>[1.00] | -2.90×10<sup>-3</sup><br>(2.81×10<sup>-3</sup>)<br>[1.00] | -5.98×10<sup>-3</sup><br>(4.78×10<sup>-3</sup>)<br>[0.99] | -1.10×10<sup>-3</sup><br>(3.47×10<sup>-3</sup>)<br>[1.00] | -4.52×10<sup>-3</sup><br>(3.58×10<sup>-3</sup>)<br>[1.00] | -3.03×10<sup>-3</sup><br>(4.48×10<sup>-3</sup>)<br>[1.00] |
| Firm size×Meat main |  |  |  | 8.60×10<sup>-4***</sup><br>(2.10×10<sup>-4</sup>)<br>[1.00] | 3.98×10<sup>-4</sup><br>(2.52×10<sup>-4</sup>)<br>[1.00] | 5.63×10<sup>-4*</sup><br>(3.43×10<sup>-4</sup>)<br>[1.00] |  |  |  |
| Lag(Recall size) |  |  |  |  |  |  | -8.50×10<sup>-6</sup><br>(2.10×10<sup>-5</sup>)<br>[1.00] | -1.71×10<sup>-5</sup><br>(1.61×10<sup>-5</sup>)<br>[1.00] | -1.43×10<sup>-5</sup><br>(2.68×10<sup>-5</sup>)<br>[1.00] |
| Lag(Class I) |  |  |  |  |  |  | 1.01<br>(0.68)<br>[2.76] | 1.14*<br>(0.67)<br>[3.13] | 0.95<br>(0.68)<br>[2.58] |
| Lag(Class II) |  |  |  |  |  |  | 1.06<br>(0.74)<br>[2.88] | 1.65**<br>(0.75)<br>[5.20] | 0.79<br>(0.72)<br>[2.20] |
| Observations | 133 | 133 | 133 | 133 | 133 | 133 | 100 | 100 | 100 |
| Pseudo R<sup>2</sup> | 0.20 | 0.10 | 0.58 | 0.28 | 0.12 | 0.57 | 0.20 | 0.19 | 0.37 |
| Likelihood ratio test | 28.93*** | 14.48*** | 116*** | 44.10*** | 16.93*** | 112.20*** | 21.90*** | 20.75*** | 46.88*** |
| Wald test | 25.86*** | 13.73*** | – | 61.63*** | 22.37*** | – | 31.98*** | 31.92*** | – |
| PH test | 0.71 | 1.59 | 3.68 | 2.86 | 2.29 | 3.17 | 11.51* | 9.51 | 8.05 |
| Variance of random effect ($\theta$) | – | – | 0.72 | – | – | 0.58 | – | – | 0.27 |

Notes: Reported are coefficient estimates ( $\beta$ 's) and exponentiated coefficient estimates (in brackets) from the AG, PWP-GT, and frailty models fitted to the first eight strata, corresponding to the mean number of recalls per firm. Robust standard errors are reported in parentheses for the AG and PWP-GT models, while conventional standard errors are reported for the frailty models. The PH test reports the chi-squared statistic for a test of the proportional hazards (PH) assumption, under the null hypothesis that the PH assumption holds. Pseudo R<sup>2</sup> is based on the Cox-Snell measure, which has an upper bound below 1. The likelihood-ratio and Wald tests assess the joint significance of the model coefficients, with the null hypothesis that $\beta_1 = \dots = \beta_p = 0$ . Statistical significance is indicated by \*p<0.1; \*\*p<0.05, \*\*\*p<0.01.

Thus, firms that have been in business longer do not necessarily have greater immunity to food safety incidents. In contrast, *Firm Size* is statistically significant across specifications and models, and is positively associated with both overall recall risk and the risk of subsequent recalls. For every $1 million increase in firm size, the overall recall hazard increases by 0.00404–0.00483% in the AG specifications and by 0.00296–0.00377% in the frailty specifications, indicating that accounting for unobserved firm heterogeneity somewhat attenuates the estimated effect of firm size. Further, the PWP-GT estimates suggest that a $1 million increase in firm size is associated with a 0.00164-0.00239% increase in the hazard of the next recall. Given the mean firm size of $6,253.06 million (Table 2), these effects can accumulate to economically meaningful differences in recall risk across firms. Overall, the results indicate that larger firms tend to experience recalls more frequently, both in terms of overall recall risk and the timing of subsequent recalls.

Similarly, *Meat Main* is statistically significant across specifications (1-3, 7-9) and models. In particular, the overall recall hazard for firms whose main output is meat and poultry products is approximately 2.92–2.95 times higher in the AG specifications and 2.83–2.93 times higher in the frailty specifications than that of more diversified firms. Firms whose main output is meat and poultry products also face a higher risk of subsequent recalls, with 2.18–2.50 times the risk of more diversified firms in the PWP specifications. This may be because meat and poultry naturally contain a greater number of pathogenic bacteria than other food categories.

Given the significant effects of firm size and diversification, a natural question arises: Does the recall risk associated with firm growth vary by the level of diversification? To address this question, specifications (4-6) in Table 3 investigate the joint effect of *Firm Size* and *Meat Main*. When the interaction term between *Firm Size* and *Meat Main* is included, it is found to be positive and statistically significant in the AG and frailty models but not in the PWP-GT model. This suggests that the higher overall recall risk associated with firm size is not uniform across firms: it is greater for firms that primarily produce meat and poultry products than for more diversified firms. The interaction coefficient estimates from the AG and frailty models indicate that, for every $1 million increase in firm size, primarily meat-producing firms (*Meat Main* = 1) experience an additional 0.0860% and 0.0563% increase in overall recall hazard, respectively, relative to more diversified firms (*Meat Main* = 0). This finding indicates that diversified firms experience a smaller increase in recall risk as they grow than firms focused primarily on meat and poultry products.

Turning to recall-specific factors, it is essential to remember that the size and class of a given recall event are not determinants of that recall but rather its consequences. Including these variables directly as covariates would therefore be inappropriate. To examine whether characteristics of a previous recall affect the risk of subsequent recalls, we lag these variables, meaning we lose the first recall event for each firm (i.e., stratum 1). Specifications (7-9) in Table 3 incorporate these lagged variables. Notably, the effects of the firm-specific covariates (*Firm Size* and *Meat Main*) remain quantitatively and qualitatively consistent with those reported in specifications (1-3).

Among the recall-specific characteristics considered, the lagged indicators for *Class I* and *Class II* are statistically significant in the PWP-GT model. Firms that previously experienced a Class II recall have a subsequent recall hazard approximately 5.20 times higher than firms that previously experienced a Class III recall (the reference category). Similarly, firms that previously experienced a Class I recall face a subsequent recall hazard approximately 3.13 times higher than firms with a prior Class III recall. Comparing the two, the estimated hazard is slightly higher for firms with a prior Class II recall than for those with a prior Class I recall. Therefore, although both Class I and Class II recalls are associated with a higher hazard of a subsequent recall than Class III recalls, the estimates suggest, at least qualitatively, that the hazard of a subsequent recall declines with the severity of the previous recall.

In terms of model diagnostics, the estimated variance of the random effects, Var(*u_i_*) = *θ*, in the frailty models ranges from 0.27 to 0.72 across specifications (3, 6, 9) in Table 3, suggesting the presence of non-negligible persistent unobserved firm-level heterogeneity. Consistent with this finding, the AG coefficients become somewhat smaller in magnitude after accounting for firm-specific frailty *u_i_*. This attenuation suggests that part of the association between the observed covariates and recall hazard in the AG models may be related to persistent unobserved firm-level heterogeneity. Finally, the PH assumption is satisfied across the specifications and models reported in Table 3.

Table 4 shows the estimation results from the analysis using the full sample. The results are qualitatively similar to those from the analysis using the first eight strata, supporting the robustness of our findings to alternative numbers of strata and, consequently, to the corresponding differences in sample size. Nonetheless, the inclusion of sparsely populated strata beyond the first eight palpably affects the precision of the estimates, as anticipated (Kelly and Lim, 2000; Kleinbaum and Klein, 2005; Amorim and Cai, 2015).^8^ A notable observation from the full sample analysis is more pronounced evidence of an association between past recall characteristics and overall and subsequent recall risks in specifications (7-8). Specifically, the AG model now yields statistically significant coefficients on the lagged indicators for *Class I* and *Class II*, while the PWP-GT model yields statistically significant coefficients on lagged *Recall Size* and recall class. The parameter estimate for *Recall Size* indicates that the hazard of a subsequent recall decreases with the quantity of meat recalled in the previous recall event: for every additional one thousand pounds of meat recalled in the previous recall event, the hazard of a firm’s next recall falls by approximately 0.00172%. Given the mean recall size of 1,669.34 thousand pounds (Table 2), this effect can accumulate to economically meaningful difference in subsequent recall risk.

**Table 4:**
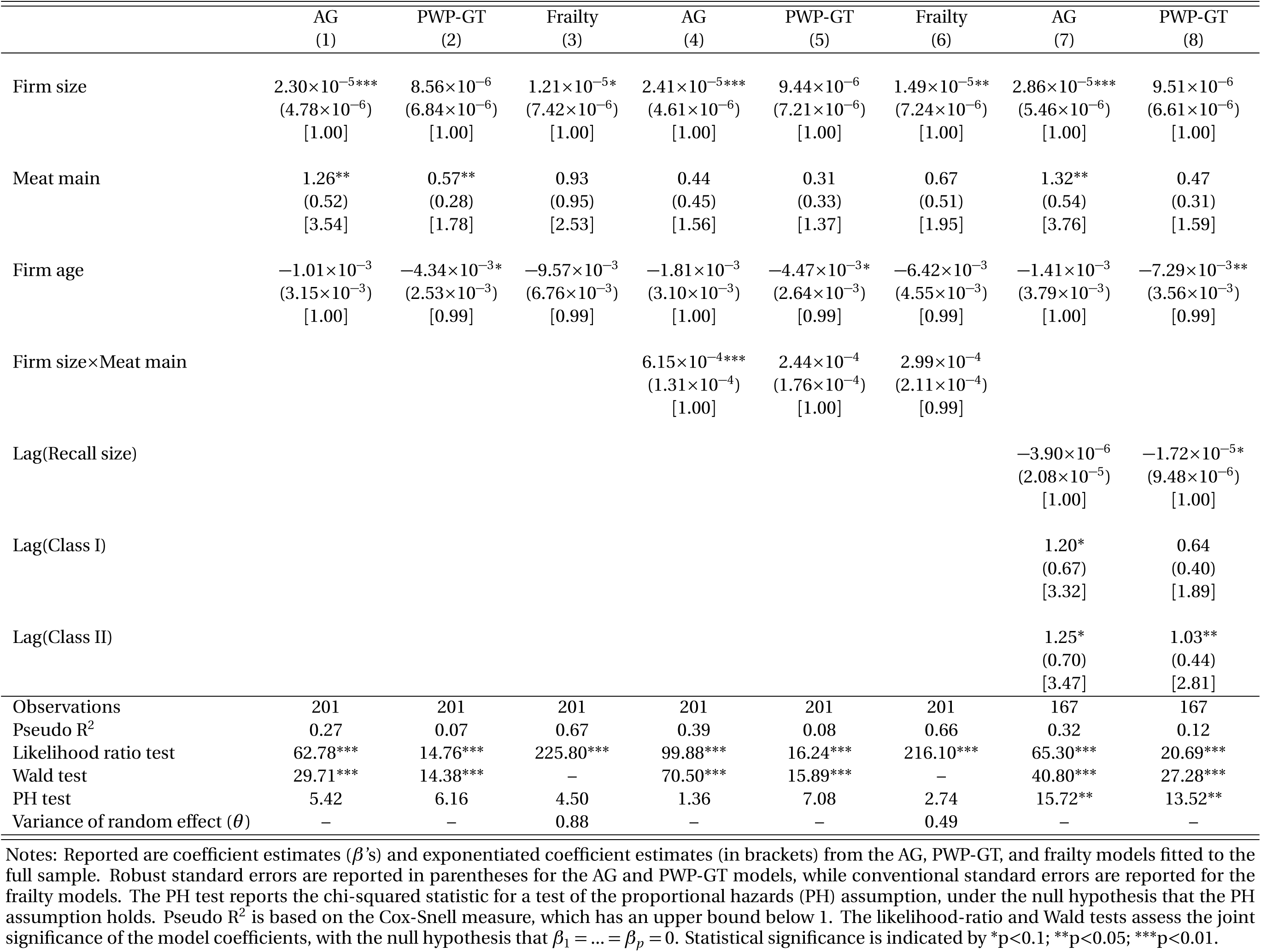
Estimates from recurrent-event survival models: full sample.

### 4.2 Decomposing Survivorship by Firm and Recall Characteristics

Figure 2 depicts the estimated survival functions for the AG and frailty models using the full sample. The survival function represents the probability of surviving (i.e., not experiencing a recall event) beyond *t* periods, i.e., Prob(*T > t*). Since the AG and frailty models focus on overall recall risk, they yield a single survival curve. The two survival curves are virtually identical, suggesting that unobserved firm-level heterogeneity is non-negligible but not substantial. The survival curves decline relatively steeply over the first 60 months compared with the period thereafter, indicating that the probability of remaining recall-free declines more rapidly during the first 60 months and then decreases at a considerably slower rate. In other words, firms are generally most prone to experiencing a recall within 60 months of operation, either from the start of the business or since the previous recall event.

**Figure 2:**
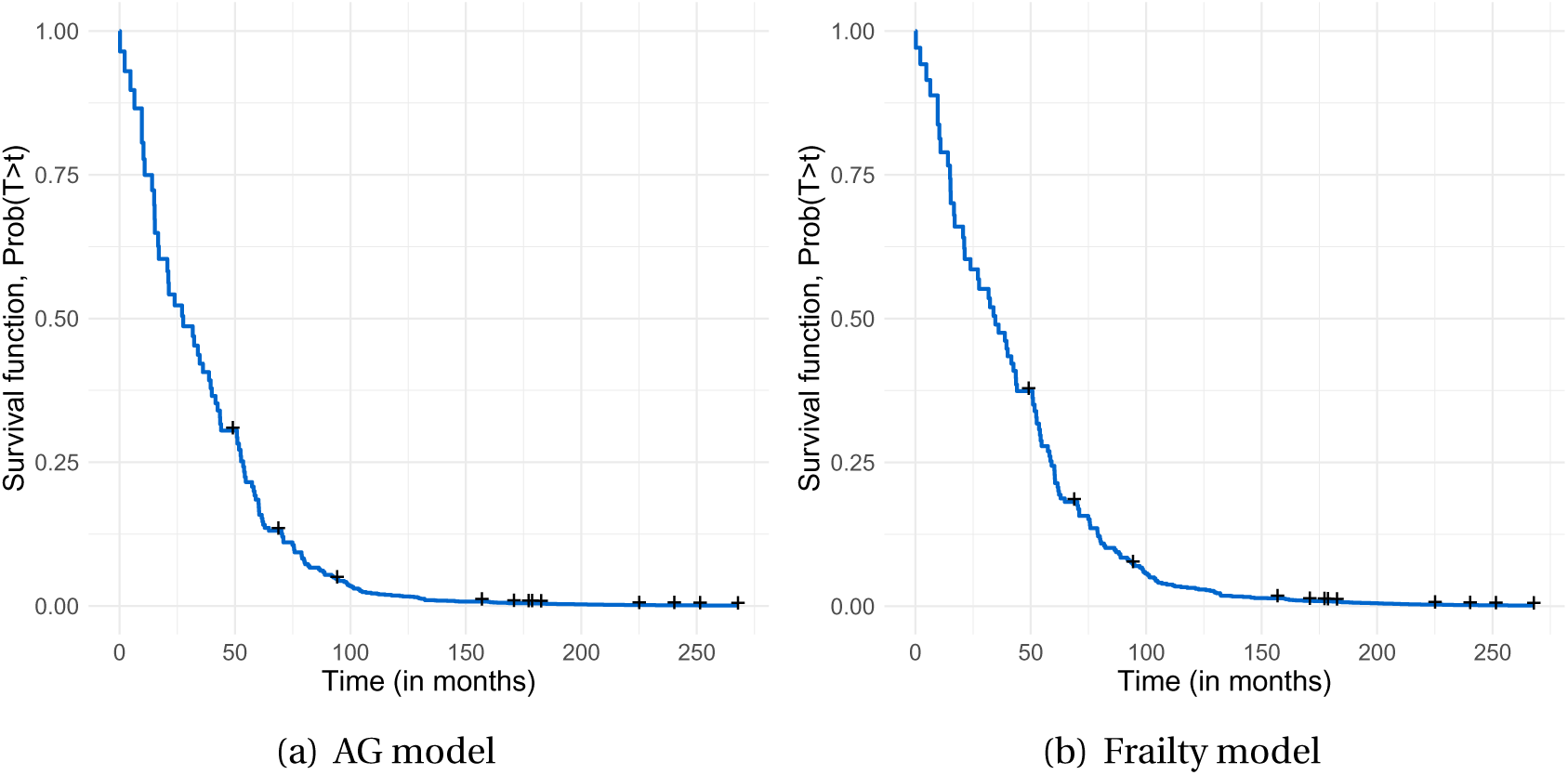
Estimated survival functions: full sample. Notes: The estimated survival function represents the probability of surviving (i.e., remaining free of a recall event) beyond *t* periods, Prob(*T > t*). Small “+” marks indicate right-censored survival times.

To illuminate the effects of firm- and recall-specific factors on the likelihood of survivorship, we next decompose the survival curves for each stratum by measurable covariates, focusing on those found to be statistically significant in Table 3, using standard Kaplan-Meier (KM) survivorship curves (Kaplan and Meier, 1958). To ensure adequate sample sizes for estimation and inferences within each stratum (Kelly and Lim, 2000; Amorim and Cai, 2015), this portion of the analysis again focuses on the first eight event strata, corresponding to the average number of recalls per firm (Table 2). Including strata beyond the eighth would increase the variance of the estimates because only a small number of firms in the data experience nine or more recall events (Table 1).

The KM survival curves are estimated non-parametrically for each stratum as:

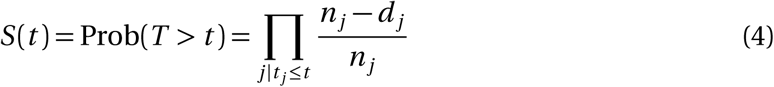

where *n_j_* is the number of observations at risk immediately prior to time *t _j_*, and *d_j_* is the number of events (recalls) occurring at time *t _j_*. Time (in months) is plotted against the estimated probability of surviving beyond time *t*, Prob(*T > t*).

Figure 3(a) illustrates the stratified KM survivorship curves decomposed by *Meat Main*, a variable that indicates whether a firm’s main output is meat and poultry or whether it has a more diversified product line. As shown, the estimated KM curves for firms whose primary output is meat and poultry consistently lie below those of diversified firms across all strata analyzed. This indicates that firms primarily producing meat and poultry have a lower probability of remaining recall-free at any given time than their more diversified counterparts. This observation is consistent with our findings from Section 4.1, but at a more granular level.

**Figure 3:**
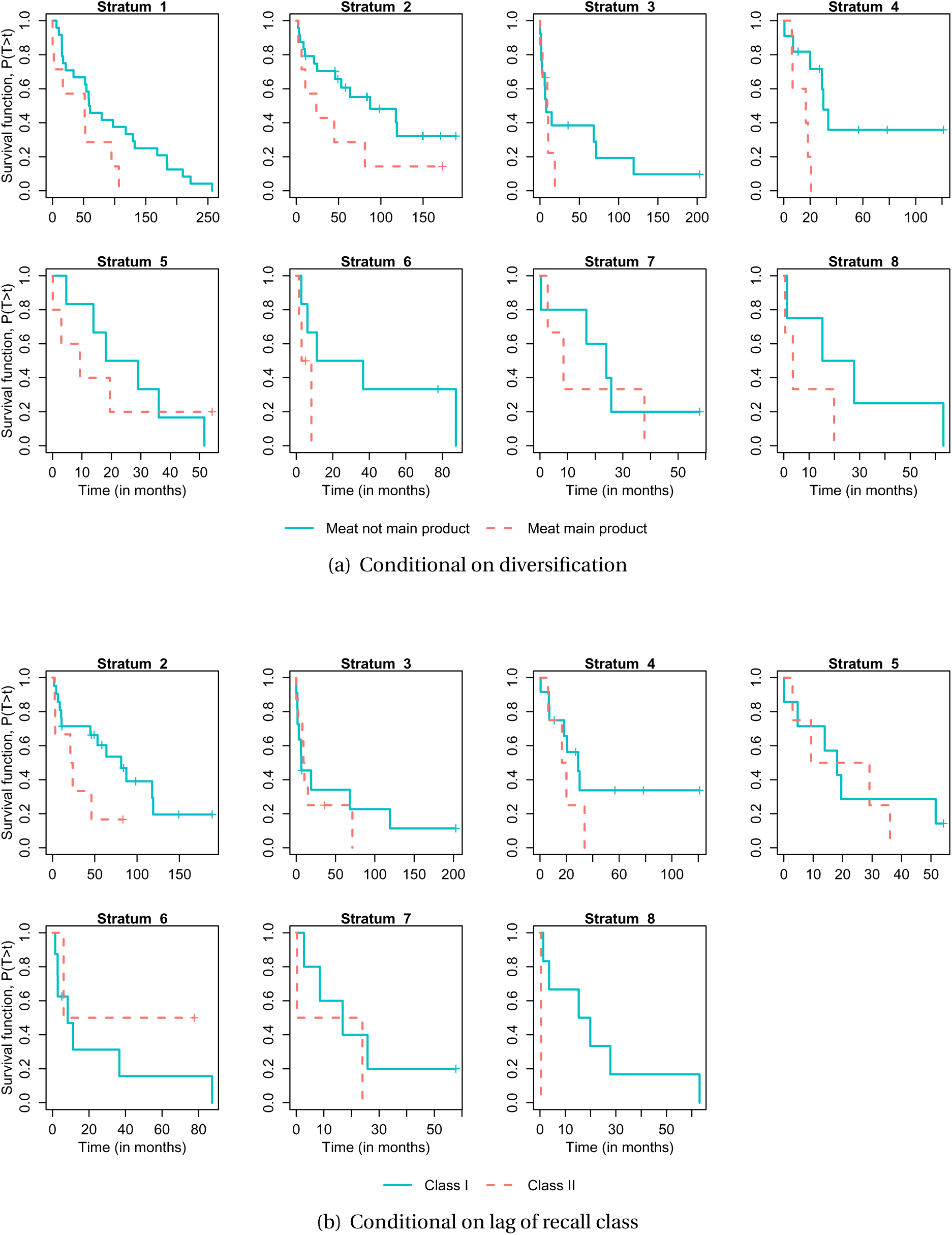
Estimated Kaplan-Meier survival functions: first eight strata. Notes: The estimated survival function represents the probability of surviving (i.e., remaining free of a recall event) beyond *t* periods, Prob(*T > t*). Small “+” marks indicate right-censored survival times.

In Figure 3(b), we decompose the survival curves by recall class. The survival curves for firms that previously experienced a Class I recall are generally higher than those for firms that previously experienced a Class II recall. This implies that the time to a subsequent recall is generally longer following a Class I recall than following a Class II recall. This observation too is in line with our findings from Section 4.1. Because Class III recalls occur only a few times in the data (Table 2), robust inference for this category is no possible; accordingly, Class III recalls are excluded from this analysis.

## 5 Conclusion

While recurrent-event survival analysis methods have been widely applied in other disciplines, their adoption in economics has been relatively limited. This study attempts to address this gap by presenting a systematic framework for analyzing recurrent time-to-event data, with the main goal of identifying factors associated with units’ risk of repeated events. We illustrate this framework using data on meat and poultry recalls issued by publicly traded firms in the United States between 1994 and 2015. We find that recall risk increases with firm size, decreases with firm diversification, and decreases with firm size among diversified firms. We also find that the risk of a subsequent recall decreases with the severity of previous recalls. These findings provide insights into the firm-level factors associated with recurrent recalls and may inform food firms’ efforts to strengthen food-safety practices and policymakers’ efforts to improve food-safety oversight. More broadly, the methodological framework presented here has fruitful applications in other areas of economics, including health economics, industrial organization, and environmental economics, where recurrent time-to-event data are commonly observed.

## Data Availability

All data used in this study are publicly available, and their sources are cited in the manuscript.

## Footnotes

1 We use “foodborne disease outbreak” and “foodborne outbreak” interchangeably throughout the article.

2 Common foodborne pathogens include Salmonella, E. coli, Listeria, Campylobacter, Clostridium perfringens, and Yersinia (CDC, 2020).

3 Not all slaughter establishments are inspected directly by the FSIS; some are inspected by state agencies. State-federal cooperative inspection programs are required by law to provide inspection that is “at least equal to” federal inspection in regulatory rigor, as mandated by the 1967 Federal Meat Inspection Act and the 1968 Wholesome Poultry Products Act.

4 In line with the existing literature, we focus on publicly traded firms because financial and accounting information is generally unavailable for privately held companies. In addition, many major meat and poultry processors and marketers are publicly traded, while privately held firms tend to be smaller (Hall and Johnson-Hall, 2017).

5 Right censoring is a standard feature of time-to-event data and is explicitly accommodated by survival models (Kleinbaum and Klein, 2005; Hosmer et al., 2011). A firm is right-censored when it leaves the risk set before experiencing the event of interest or remains event-free through the end of the observation period. Censored observations remain informative because they contribute to the risk set up to the point of censoring.

6 The CPI data are available from the U.S. Bureau of Labor Statistics at https://data.bls.gov/cgi-bin/surveymost?cu.

7 Diversified firms may produce numerous products, making product- or segment-specific production levels difficult to identify at the time of a recall. Firms also generally do not disclose expenditures on food-safety technologies, sanitation protocols, or related investments.

8 This also explains why the frailty model incorporating recall characteristics is not estimable due to convergence issues in Table 4, unlike in Table 3.

## References

Akhundjanov, S. B., Pozo, V. F., and Thomas, B. (2024). Inference on learning upon repeated compliance issues. Economics Letters, 242:111871.

Amorim, L. D. A. F. and Cai, J. (2015). Modelling recurrent events: A tutorial for analysis in epidemiology. International Journal of Epidemiology, 44(1):324–333.

Andersen, P. K. and Gill, R. D. (1982). Cox’s regression model for counting processes: A large sample study. The Annals of Statistics, 10(4):1100–1120.

Balan, T. A. and Putter, H. (2020). A tutorial on frailty models. Statistical Methods in Medical Research, 29(11):3424–3454.

Breslow, N. (1974). Covariance analysis of censored survival data. Biometrics, 30(1):89–99.

CDC (2020). Foods that can cause food poisoning. Retrieved from: https://www.cdc.gov/foodsafety/foods-linked-illness.html.

Chen, Y., Ganesan, S., and Liu, Y. (2009). Does a firm’s product-recall strategy affect its financial value? An examination of strategic alternatives during product-harm crises. Journal of Marketing, 73(6):214–226.

Cox, D. R. (1972). Regression models and life tables. Journal of the Royal Statistical Society: Series B, 34(2):187–220.

Duchateau, L., Janssen, P., Kezic, I., and Fortpied, C. (2003). Evolution of recurrent asthma event rate over time in frailty models. Journal of the Royal Statistical Society: Series C, 52(3):355–363.

Fama, E. and French, K. (1992). The cross-section of expected stock returns. Journal of Finance, 47(2):427–465.

FDA (2017). Recalls of foods and dietary supplements. Retrieved from: https://www.fda.gov/Food/RecallsOutbreaksEmergencies/Recalls/default.htm.

FSIS (2013). Recall of meat and poultry products. FSIS Directive 8080.1, Washington DC.

FSIS (2017). Recall case archive. Retrieved from: https://www.fsis.usda.gov/wps/portal/fsis/topics/recalls-and-public-health-alerts/recall-case-archive.

Grambsch, P. M. and Therneau, T. M. (1994). Proportional hazards tests and diagnostics based on weighted residuals. Biometrika, 81(3):515–526.

Hall, D. C. and Johnson-Hall, T. D. (2017). Learning from conformance quality failures that triggered product recalls: The role of direct and indirect experience. Journal of Supply Chain Management, 53(4):13–36.

Henson, S. and Mazzocchi, M. (2002). Impact of bovine spongiform encephalopathy on agribusiness in the United Kingdom: Results of an event study of equity prices. American Journal of Agricultural Economics, 84(2):370–386.

Henson, S. and Reardon, T. (2005). Private agri-food standards: Implications for food policy and the agri-food system. Food Policy, 30(3):241–253.

Hosmer, D. W., Lemeshow, S., and May, S. (2011). Applied Survival Analysis: Regression Modeling of Time-to-Event Data. John Wiley & Sons, New York.

Hua, X. (2011). Product recall and liability. Journal of Law, Economics, & Organization, 27(1):113–136.

Jarrell, G. and Peltzman, S. (1985). The impact of product recalls on the wealth of sellers. Journal of Political Economy, 93(3):512–536.

Kaplan, E. L. and Meier, P. (1958). Nonparametric estimation from incomplete observations. Journal of the American Statistical Association, 53(282):457–481.

Kelly, P. J. and Lim, L. L.-Y. (2000). Survival analysis for recurrent event data: An application to childhood infectious diseases. Statistics in Medicine, 19(1):13–33.

Kennedy, B., Kasl, S., and Vaccarino, V. (2001). Repeated hospitalizations and self-rated health among the elderly: A multivariate failure time analysis. American Journal of Epidemiology, 153(3):232–241.

Kleinbaum, D. G. and Klein, M. (2005). Survival Analysis: A Self-Learning Text. Springer, New York, 2 edition.

Marsh, T. L., Schroeder, T. C., and Mintert, J. (2004). Impacts of meat product recalls on consumer demand in the USA. Applied Economics, 36(9):897–909.

Mazzocchi, M. (2006). No news is good news: Stochastic parameters versus media coverage indices in demand models after food scares. American Journal of Agricultural Economics, 88(3):727–741.

McCluskey, J. J., Grimsrud, K. M., Ouchi, H., and Wahl, T. I. (2005). Bovine spongiform encephalopathy in Japan: Consumers’ food safety perceptions and willingness to pay for tested beef. Australian Journal of Agricultural and Resource Economics, 49(2):197–209.

Ollinger, M., Moore, D. L., and Chandran, R. (2004). Meat and poultry plants’ food safety investments: Survey findings. Technical Bulletin No. 1911. Washington DC: Economic Research Service.

Ollinger, M. and Mueller, V. (2003). Managing for safer food: The economics of sanitation and process controls in meat and poultry establishments. Agricultural Economics Report No. 817. Washington DC: Economic Research Service.

Piggott, N. E. and Marsh, T. L. (2004). Does food safety information impact US meat demand? American Journal of Agricultural Economics, 86(1):154–174.

Prentice, R. L., Williams, B. J., and Peterson, A. V. (1981). On the regression analysis of multivariate failure time data. Biometrika, 68(2):373–379.

Pruitt, S. W. and Peterson, D. R. (1986). Security price reactions around product recall announcements. Journal of Financial Research, 9(2):113–122.

Rhee, M. and Haunschild, P. R. (2006). The liability of good reputation: A study of product recalls in the US automobile industry. Organization Science, 17(1):101–117.

Rondeau, V., Mathoulin-Pelissier, S., Jacqmin-Gadda, H., Brouste, V., and Soubeyran, P. (2007). Joint frailty models for recurring events and death using maximum penalized likelihood estimation: Application on cancer events. Biostatistics, 8(4):708–721.

Schmall, E. (2024). Is the food supply less safe than it used to be? The New York Times. Retrieved from: https://www.nytimes.com/2024/11/25/well/food-safety-outbreaks.html.

Schroeder, T. C., Tonsor, G. T., Pennings, J. M. E., and Mintert, J. (2007). Consumer food safety risk perceptions and attitudes: Impacts on beef consumption across countries. The B.E. Journal of Economic Analysis & Policy, 7(1).

Sockett, P. (1993). Social and economic aspects of food-borne disease. Food Policy, 18(2):110–119.

Spalding, A., Goodhue, R. E., Kiesel, K., and Sexton, R. J. (2023). Economic impacts of food safety incidents in a modern supply chain: E. coli in the romaine lettuce industry. American Journal of Agricultural Economics, 105(2):597–623.

Sultana, N., Alam, M., and Hasinur Rahaman Khan, M. (2026). Joint frailty mixture cure model for recurrent event data with dependent censoring: An MCEM approach. Statistics in Medicine, 45(10-12):e70579.

Teratanavat, R. and Hooker, N. H. (2004). Understanding the characteristics of US meat and poultry recalls: 1994–2002. Food Control, 15(5):359–367.

Teratanavat, R., Salin, V., and Hooker, N. (2005). Recall event timing: Measures of managerial performance in U.S. meat and poultry plants. Agribusiness, 21(3):351–373.

Welling, L. (1991). A theory of voluntary recalls and product liability. Southern Economic Journal, 57(4):1092–1111.

